# Circulating-tumor DNA from Two Types of Blood Collection Tubes for Monitoring Tumor Burden Validated with Esophageal Squamous Cell Carcinoma Patients

**DOI:** 10.1101/2020.08.12.20173070

**Authors:** Fumitaka Endo, Takeshi Iwaya, Yasushi Sasaki, Takehiro Chiba, Mizunori Yaegashi, Kohei Kume, Kei Sato, Atsuhiro Arisue, Ryoko Kawagishi, Teppei Matuo, Yuji Akiyama, Akira Sasaki, Yuji Suzuki, Takashi Tokino, Mari Masuda, Tesshi Yamada, Hidewaki Nakagawa, Satoshi S. Nishizuka

## Abstract

**Objective:** To evaluate long-term whole blood (WB) storage at room temperature (RT), plasma and peripheral blood mononuclear cell (PBMC) DNA was collected simultaneously using BD Vacutainer^®^ CPT^TM^ Mononuclear Cell Preparation Tubes (CPT) and Streck Cell-Free DNA Blood Collection Tubes (BCT^®^).

**Methods:** Plasma DNA was isolated from both types of tubes at various time points at RT. DNA from PBMCs was extracted using CPT from WB stored in BCT. The extracted DNA was used to monitor esophageal cancer treatment.

**Results:** BCT maintained steady levels of plasma DNA for up to nine days. After transfer of BCT-stored WB to CPT, PBMC-DNA were yielded with suitable quality for up to seven days. ctDNA from esophageal cancer patients carrying *TP53* mutations reflected treatment efficacy.

**Conclusion:** BCT/CPT combinatory procedure allows storage of blood samples for up to seven days at RT for valid clinical assays using ctDNA.

---

Circulating-tumor DNA (ctDNA) is one of the most promising new classes of biomarkers in the field of clinical oncology. The bloodstream of cancer patients carries an extremely small fraction of ctDNA in fragmented plasma genomic DNA (i.e., cell-free DNA) that is released from various types of nucleated cells in the body.^1, 2^ Thus, to quantitatively monitor ctDNA in clinical practice, it is necessary to obtain the appropriate blood fraction with a simple and reproducible procedure. However, conventional blood collection tubes (e.g., K_3_EDTA tubes) generally require prompt centrifugation (e.g., within two hours) to separate plasma or peripheral blood mononuclear cell (PBMC) layers as these tubes do not contain fixatives.^3-5^ This time-sensitive centrifuge step and need for layer separation represents a major obstacle for widespread use of ctDNA as a diagnostic biomarker for tumor burden monitoring.

BD Vacutainer^®^ CPT™ Mononuclear Cell Preparation Tubes (CPT) are widely used to simultaneously separate plasma and PBMCs. CPT contain sodium citrate as an anticoagulant, a density-gradient FICOLL™ Hypaque™, and a polyester gel barrier.^6, 7^ Centrifugation of samples in CPT within two hours of whole blood collection readily isolates a visibly distinct plasma fraction and a PBMC layer (i.e., buffy coat). The resulting plasma and PBMCs can be stored at −80 °C in the respective separate tubes. The two-hour time restriction is imposed to avoid cellular DNA release from nucleated cells into plasma by cytolysis. Such cytolytic release of cellular DNA can reduce the proportion of ctDNA in plasma DNA, resulting in an artifact of an extremely low variant allele frequency (VAF). In our previous study, the ctDNA VAF was very low (ranging from 0 to 1.12%) and may be susceptible to DNA released from nucleated cells during quantitative analysis in preoperative colorectal cancer patients.^8^ Therefore, the need to centrifuge CPT in order to obtain ctDNA and PBMCs compatible for quantitative analysis could limit the information gained from ctDNA. In contrast, Cell-Free DNA Blood Collection Tubes (BCT^®^) have been commercialized by Streck (La Vista, NE) and allow for the isolation of plasma DNA after storage of samples between 6 °C and 37 °C for up to 14 days after sample collection.^3-5^ The fixative in BCT prevents DNA release from nucleated cells into the plasma thus providing a longer time window for isolation of high-quality ctDNA. BCT partition PBMCs into the pale layer boundary between plasma and a lower layer containing RBC during the first centrifuge, but PBMCs cannot be collected separately from RBC. However, previous reports have suggested that the integrity of nucleated cells in peripheral blood was maintained for at least 4 days.^9^ Overall, the characteristics of BCT offer an opportunity to maintain whole blood for ctDNA isolation without additional introduction of cellular DNA and the subsequent extraction of DNA from PBMCs without cytolysis.

In this study, we attempted to establish a practical procedure for tumor burden monitoring with ctDNA using CPT and BCT. For clinical practice, an acceptable whole blood (WB) preservation time window for downstream DNA extraction from both plasma and PBMCs was determined. In addition, we performed tumor burden monitoring with ctDNA for esophageal squamous cell carcinoma (ESCC) cases treated with either surgery or chemotherapy, which were both aimed at reducing tumor volume.

## Materials and Methods

### Recruitment of Healthy Blood Donors

Volunteer blood donors were recruited from Iwate Medical University employees after providing written informed consent. The donor pool included both males and females who were all presumed to be healthy. This study was approved by the Institutional Review Board of Iwate Medical University School of Medicine, Iwate, Japan (IRB: H27-55).

### Blood Collection and Plasma Processing

The study flow is illustrated in Figure 1. Briefly, a total of 96 ml of whole venous blood was collected from each of seven healthy volunteers, with eight ml collected into six CPT tubes and six BCT tubes for time-course studies. Blood samples in each tube were inverted 10 times and stored at room temperature until plasma isolation on day zero (immediately processed), three, six, nine, 12, and 14 after blood collection. Prior to the plasma separation at each time point, the tubes were again inverted 10 times and centrifuged at 1800 ×*g* for 20 min at room temperature using a swing-out rotor (KUBOTA, Tokyo, Japan). The plasma layer from CPT and BCT was carefully collected without disturbing the buffy coat and red blood cell (RBC) layer, transferred into a new 15 ml collection tube, and the plasma layer was centrifuged at room temperature at 1800 ×*g* for 20 min to remove any residual cellular debris. The supernatant (i.e., plasma) was transferred again to a new cryotube and frozen at −80 °C until DNA extraction.

**Figure 1.**
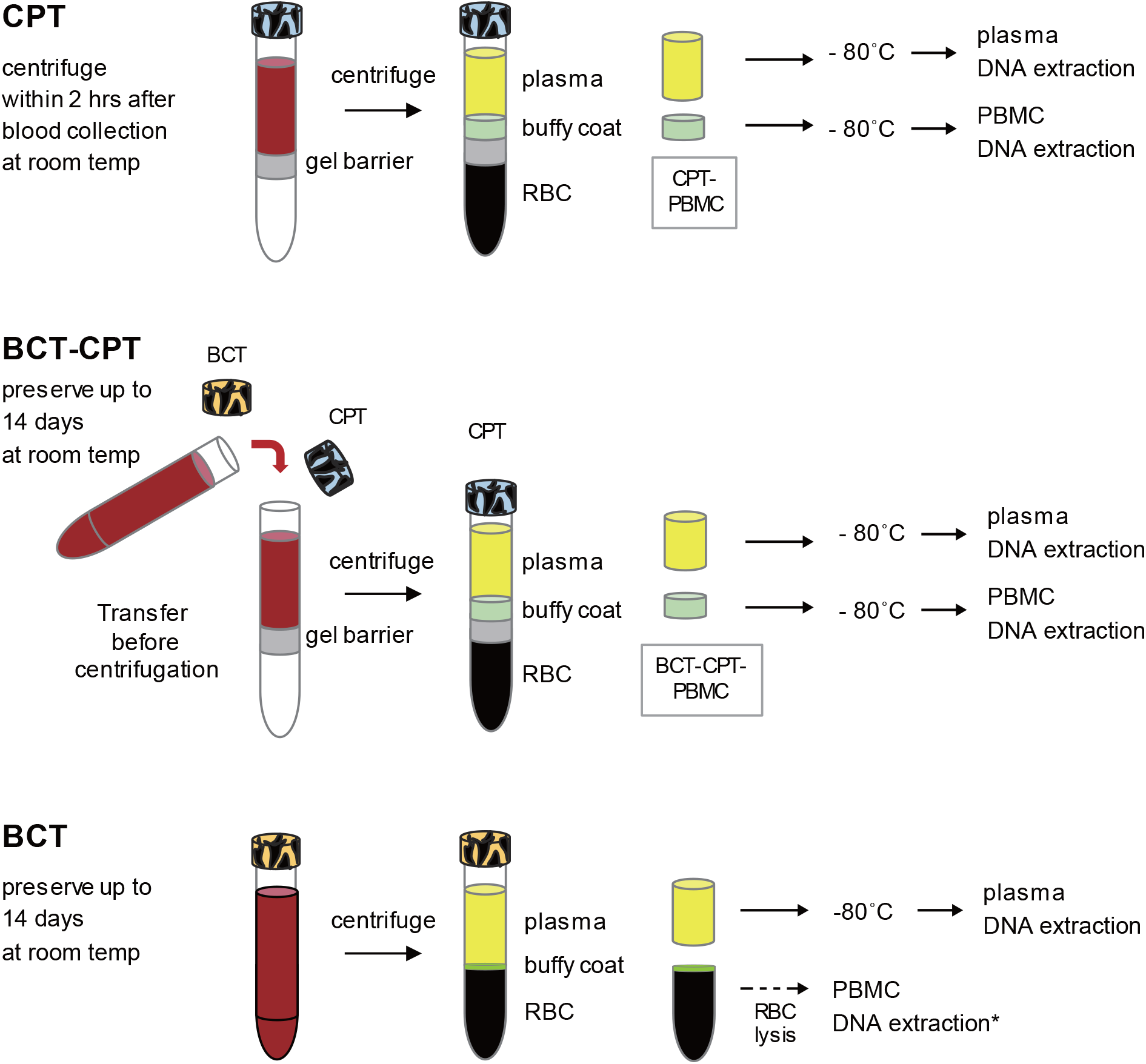
An illustration of blood sample collection processes. Bottom: BCT tube. Top: CPT tube. Middle: combinatory tube method.. CPT, Cell Preparation Tubes. BCT, Cell-Free DNA Blood Collection Tubes. RBC, red blood cell. PBMC, peripheral blood mononuclear cell.

### DNA Extraction from Plasma and PBMCs

Plasma DNA was extracted from four ml frozen plasma samples using a QIAamp^®^ Circulating Nucleic Acid Kit according to the manufacturer’s instructions (Qiagen, Hilden, Germany). Extracted DNA was eluted in 100 μl Qiagen Buffer AVE and stored at −20 °C until analysis. PBMCs were isolated at room temperature either by: (i) directly collecting the buffy coat drawn from WB in a CPT (Figure 1, top); or (ii) collecting the buffy coat from WB in BCT and then transferred into a CPT (Figure 1, middle). Collected PBMCs were transferred to a new 1.5 ml centrifuge tube and stored at −80 °C. Cellular DNA was extracted from the collected PBMCs using a QIAamp^®^ DNA Mini Kit (Qiagen, Hilden, Germany) and stored at −20 °C until analysis. The PBMC DNA yield was compared between methods (i) and (ii) at day zero and seven.

### Quantifying DNA Concentrations

The purified plasma DNA and PBMC DNA concentrations were quantified using a Qubit dsDNA HS Assay Kit according to the manufacturer’s instructions (Thermo Fisher Scientific, Waltham, MA). The genome copy number was estimated from quantitative PCR (qPCR) of the human long interspersed nuclear element-1 *(LINE-1)* gene using a LightCycler 480 Probes Master Kit (Roche Applied Science, Mannheim, Germany) according to the manufacturer’s protocol with specific primers and universal probes that were designed at the assay design center of the Universal Probe Library (https://qpcr.probefinder.com/organism.jsp). *LINE-1* primer sequences and universal probe number were: Forward, 5'-tgggggtcacatgtaatgaa-3'; reverse, 5'-gccagcttgtgtgtcattt-3'; and probe #8 (Roche Applied Science, Mannheim, Germany).

### Esophageal Cancer Patients and Sample Collection

Primary tumor samples were either surgically or endoscopically obtained from five ESCC patients. Tumor tissues were immediately cut and stored at −80 °C until DNA extraction. Corresponding blood samples were also obtained pre- and post-treatment. Based on the result from healthy volunteers, plasma and PBMC from pre-treatment blood samples were separately collected using BCT and CPT. In post-treatment blood, the plasma samples were directly collected from BCT after centrifugation (Figure 1, bottom). All plasma and PBMC samples were collected within 5 days after blood collection and stored at −80 °C until DNA extraction. All DNA samples were stored at −20 °C until analysis. The study in which ESCC patients were enrolled was approved by the IRB of Iwate Medical University (IRB# HGH27-16) and was registered in the UMIN Clinical Trial Registry (UMIN000038724).

### Amplicon Sequencing

For mutation screening of ESCC samples, five pairs of tumor tissues and corresponding PBMCs were subjected to amplicon sequencing using an Ion AmpliSeq™ *TP53* Panel, which covers all coding exons of the *TP53* gene with 24 amplicons, according to the manufacturer’s protocol (Thermo Fisher Scientific). Approximately 20 ng DNA per sample was used to prepare barcoded libraries with IonXpress barcoded adapters and the Ion AmpliSeq Library Kit 2.0 (Thermo Fisher Scientific). Ten individual barcoded libraries (100 pM each) were pooled and clonally amplified through emulsion PCR using the One Touch Instrument and the Ion PGM Template OT2 200 kit (Thermo Fisher Scientific). Finally, the sequencing of templates was performed on a PGM 316 chip using the Ion PGM 200 Sequencing Kit v2 according to the manufacturer’s instructions (200 bp read length, Thermo Fisher Scientific).

### Identification of Somatic Mutation

Genome Reference Consortium Human Build 37 (GRCh37/hg19) was used as a reference. Alignment to the GRCh37/hg19 genome and sequencing read counting were performed in Torrent Suite version 5.0 (Thermo Fisher Scientific). Somatic mutations, including single-nucleotide variants, insertions, and deletions, were detected using statistics in tumor and matched PBMC samples from the Ion Reporter software 5.0 tumor-normal workflow (Thermo Fisher Scientific), in which germline variants were subtracted from the tumor variants, as previously described.^23^ All identified single-nucleotide variants, insertions, and deletions were visually inspected using the Integrative Genomics Viewer software to filter out possible strand-specific errors, such as a mutation detected only in either the forward or reverse DNA strand. The dbSNP database was used to exclude single nucleotide polymorphisms (SNPs) from the called variants. The following criteria were used as cutoffs: total coverage >20, variant coverage >10, and variant frequency >5%. Mutations were called if they occurred in < 0.1% of reads in the normal control (minor allele frequency) and were absent from dbSNP as well as the 1000 Genomes Project database.

### dPCR

dPCR was performed with the QuantStudio 3D dPCR System (Thermo Fisher Scientific). Case-specific mutation primer/probes for dPCR were designed using the Hypercool Primer & Probe™ method (Nihon Gene Research Laboratories, Inc., Sendai, Japan). In brief, dPCR reaction mixtures contained 7.5 μl QuantStudio 3D dPCR Master Mix v2, 1.5 μl Taqman primer/probe mix, and up to 6 μl cfDNA with PCR-grade H_2_O added to yield a total volume of 15 μl. Using the QuantStudio 3D dPCR Chip Loader, samples were partitioned onto a 20,000-well QuantStudio 3D dPCR Chip v2, which was analyzed by PCR using a ProFlex 2x Flat PCR System with the following program: 10 min at 96 °C, 39 cycles of 2 min at 60 °C, followed by 30 s at 98 °C, 2 min at 60 °C, and holding at 10 °C. The dPCR data were acquired with a QuantStudio 3D dPCR instrument, and the data were analyzed with the QuantStudio 3D Analysis Suite (Version 3.0).

### Statistical Analysis

Student’s *t*-test was used to determine whether numerical values between two groups were significantly different. The *t*-test was two-tailed and a *P* < 0.05 was considered statistically significant. Measured sample values were expressed as a standard error of the mean unless otherwise noted. Pearson’s correlation coefficient (r) was used for correlation measurements between two variables. Calculations were performed using JMP 9 software (SAS Institute, Cary, NC).

## Results

### Visible hemolysis of Plasma in Both Blood Collection Tubes

After centrifugation of samples in both tube types immediately after blood collection, yellowish clear plasma and cellular layers were visible in CPT and BCT (Figure 2A, day zero). By day six, the boundary between the upper and buffy coat layers was less defined in both CPT and BCT tubes (Figure 2A, day six), and the plasma color turned a murky red (Figure 2B). These characteristics are indicative of hemolysis, but may not directly reflect DNA release from nucleated cells, particularly for BCT, which includes fixatives designed to preserve cell membranes of nucleated cells.^24^

**Figure 2.**
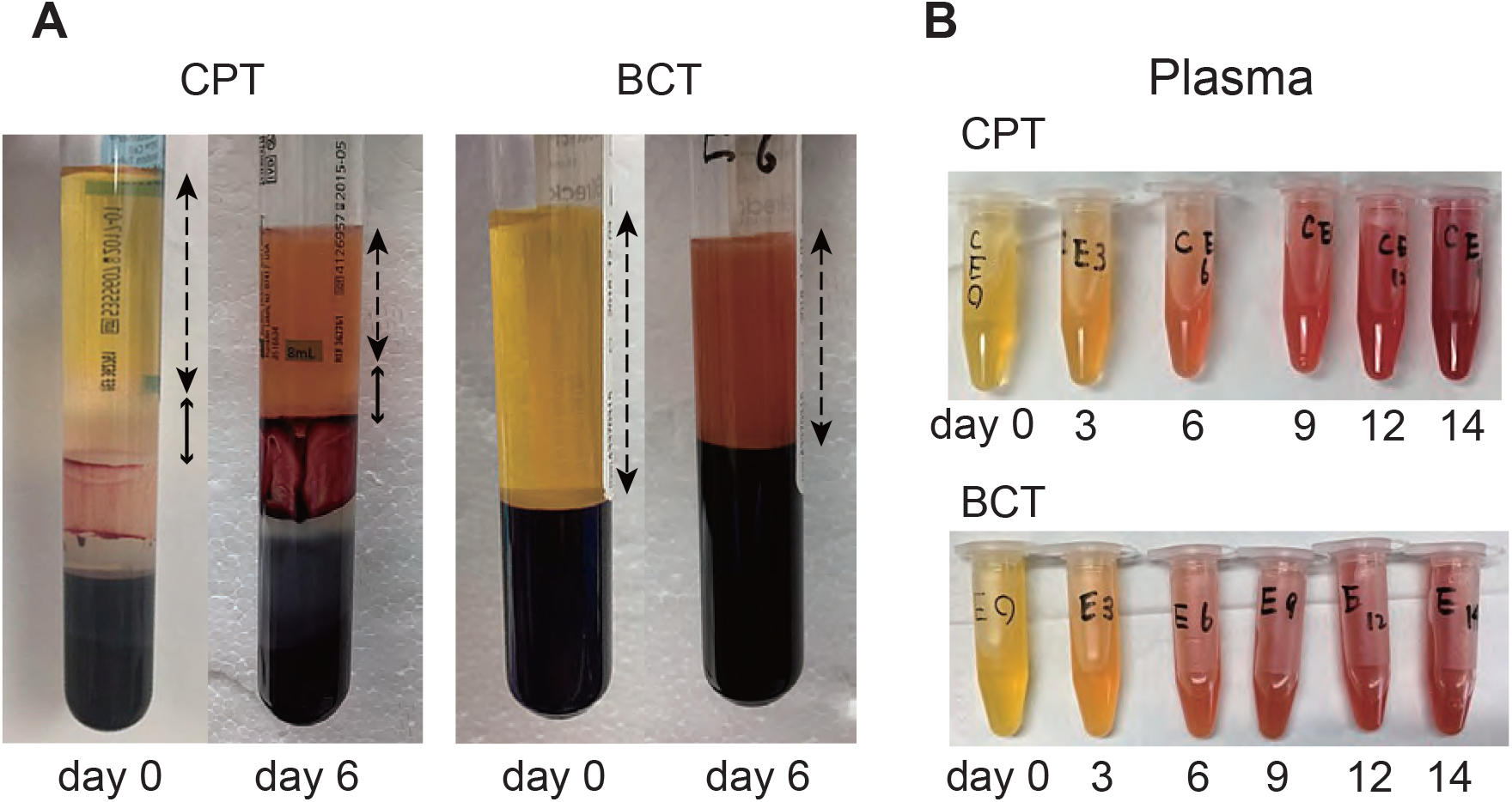
Appearance of plasma and buffy coat layers in CPT and BCT tubes after centrifugation. **A**, Comparison between day 0 and day 6 in CPT and BCT tubes. **B**, Visible cytolysis in both CPT and BCT tubes up to 14 days. Solid and dashed arrows indicate buffy coat layer and plasma layer, respectively. CPT, Cell Preparation Tubes. BCT, Cell-Free DNA Blood Collection Tubes.

### Time-Course Evaluation of DNA Levels in BCT and CPT

Fluorometry in the Qubit dsDNA HS Assay Kit (Thermo Fisher Scientific) showed that, relative to day zero, the plasma DNA level in CPT continuously increased, from 7.4-fold at day three to 237.4-fold at day 14 (Figure 3A). Furthermore, the plasma DNA level of BCT samples was significantly and consistently lower than that of CPT (3.4- and 53.1-fold at day three and day 14, respectively) (Figure 3A). The plasma DNA level of BCT samples showed no significant increases until day 14, when the maximum mean concentration was 44.7 ng/ml (Figure 3B). Although the mean level of plasma DNA of BCT did gradually increase, the change was minimal relative to that of CPT (2.2-fold vs. 229.2-fold increase on day 12 relative to day zero for BCT and CPT, respectively) (Figure 3A, B).

**Figure 3.**
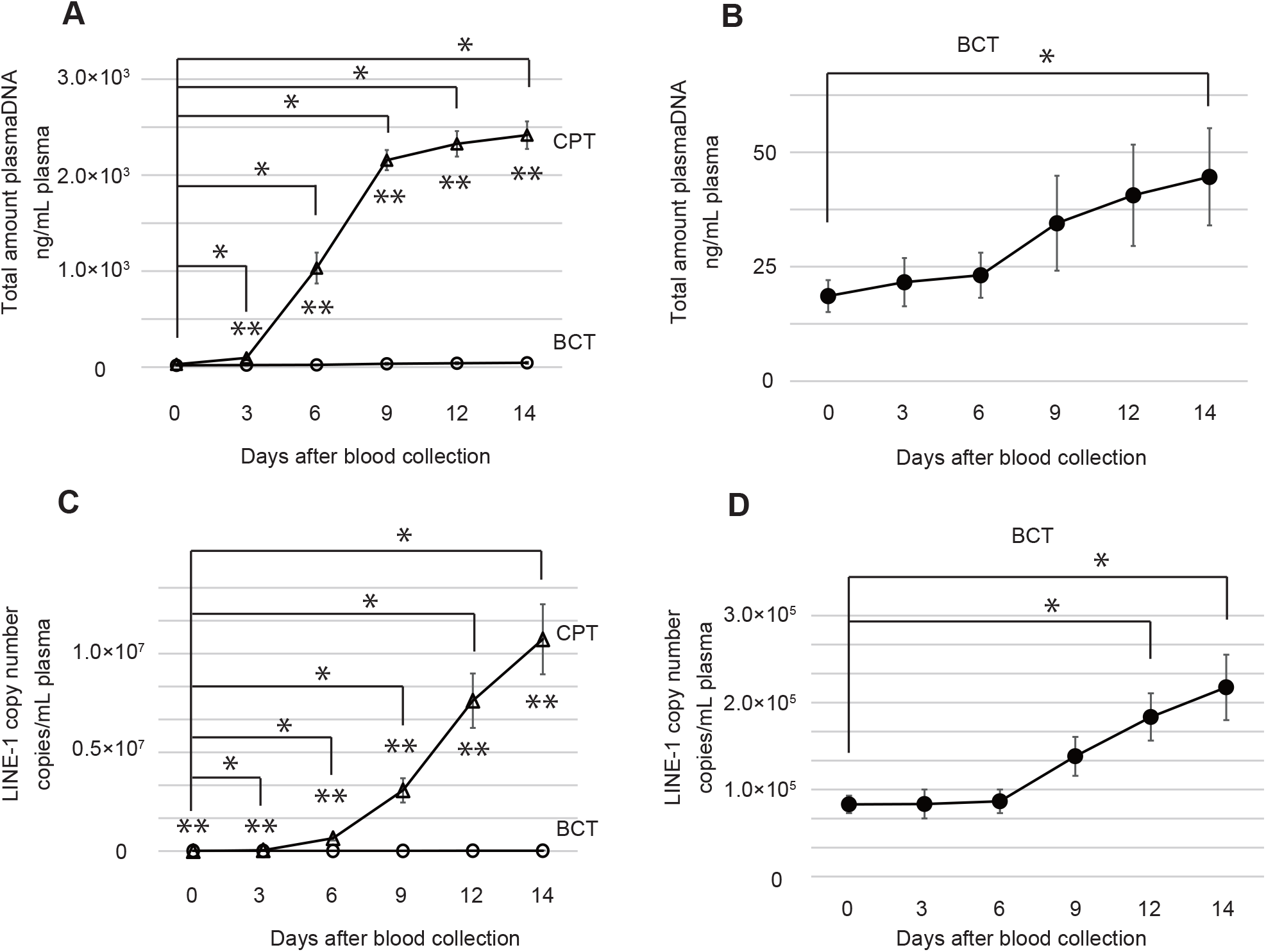
Chronological changes in cfDNA concentration. **A**, cfDNA levels in CPT and BCT. Significant differences relative to day zero in CPT are indicated by single asterisks (*t*-test, *p* < 0.05). Significant differences between CPT and BCT are indicated by double asterisks (*t*-test, *p* < 0.05). **B**, cfDNA levels in BCT. Significant differences relative to day zero are indicated by a single asterisk (*t*-test, *p* < 0.05). C, LINE-1 gene copy number in CPT and BCT. Significant differences relative to day zero in CPT are indicated by single asterisks (*t*-test, *p* < 0.05). Significant differences between CPT and BCT are indicated by double asterisks (*t*-test, *p* < 0.05). D, LINE-1 gene copy number in BCT. Significant differences relative to day zero are indicated by a single asterisk (*t*-test, *p* < 0.05). Triangles and circles indicate mean of cfDNA levels in CPT and BCT, respectively. Error bars represent ± standard error of the mean for each side. cfDNA, cell-free DNA. CPT, Cell Preparation Tubes. BCT, Cell-Free DNA Blood Collection Tubes. LINE-1, human long interspersed nuclear element-1.

The *LINE-1* retrotransposon family members comprise a significant portion of the human genome and thus allow estimation of the genome copy number.^25^ To evaluate changes in genome copy number in CPT and BCT samples, the *LINE-1* copy number was estimated by qPCR. As expected, the *LINE-1* level of both CPT and BCT showed trends that were similar to those seen for plasma DNA. Relative to day zero, the *LINE-1* level in CPT increased with time from 9.2-fold on day three to 2331.9-fold by day 14 (Figure 3C). Increases in *LINE-1* levels in BCT were significantly lower than CPT (4.8-fold on day three to 495.1-fold on day 14) (Figure 3C). Although an increasing trend was seen, the *LINE-1* level of BCT showed no significant increase until day 12, when a 2.2-fold increase relative to day zero was seen (Figure 3D). Similar to the plasma DNA level, this increase was negligible compared to the 415.4-fold increase on day 12 seen for CPT (Figure 3B).

Plasma DNA levels and genome copy number as estimated by *LINE-1* measurements were very well correlated *(r* = 0.98, *p* = 0.0004), which is consistent with our previous data^8^. Thus, both plasma DNA measurements and *LINE-1* copy number estimates suggest that ctDNA can be quantitatively measured in samples stored in BCT for up to at least nine days in the absence of significant contaminating DNA derived from nucleated cells.

### PBMC Collection from BCT with CPT

In CPT, the buffy coat containing PBMCs is a visible whitish layer that lies just above the CPT gel barrier after centrifugation. Although the buffy coat is also recognized as a pale layer in BCT, PBMCs cannot be separated from the lower layer containing RBC. No significant difference in DNA yield was observed between CPT-PBMC (PBMCs directly isolated from WB in CPT, Figure 1, top) and BCT-CPT-PBMC (PBMCs isolated from BCT-preserved WB that was transferred to CPT, Figure 1, middle). The DNA yield on day zero was 219.6 ± 12.8 and 171.7 ± 22.8 ng/ml from CPT-PBMC and BCT-CPT-PBMC, respectively (Figure 4; *p* = 0.14). Moreover, no significant difference in DNA yield from BCT-CPT-PBMC was observed between day zero and day seven (171.7 ± 22.8 ng/ml and 210.1 ± 66.5 ng/ml of WB, respectively; Figure 4; *p* = 0.61). Therefore, with the use of BCT for initial storage and transfer of samples to CPT, both plasma DNA and PBMCs could be stably collected from BCT-preserved whole blood samples for at least seven days after blood collection.

**Figure 4.**
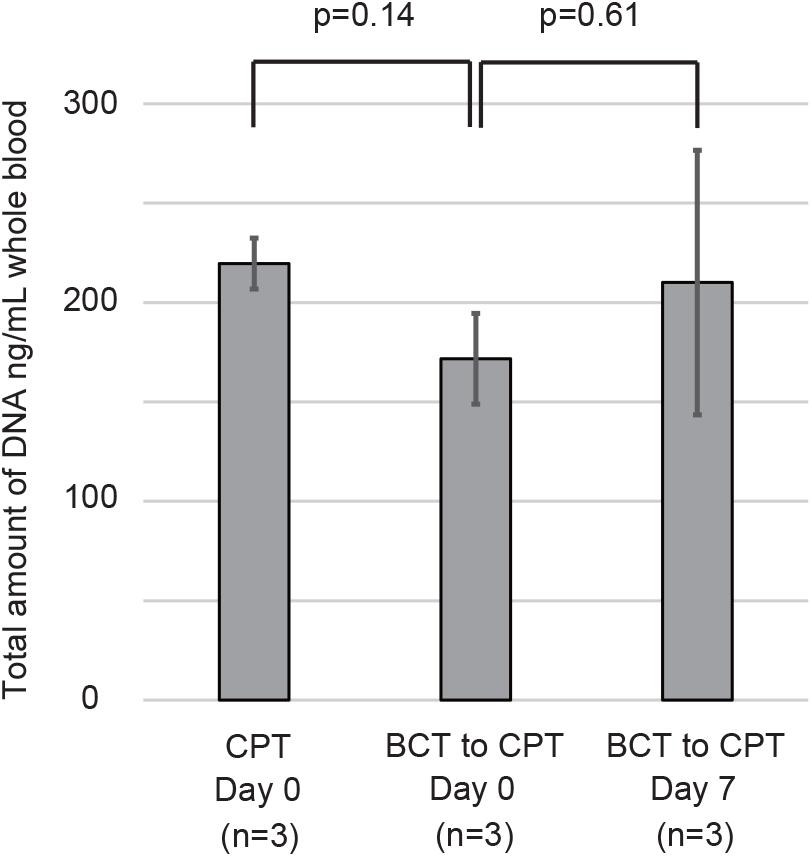
DNA yield from PBMCs isolated from whole blood samples preserved in CPT and BCT. CPT, PBMCs directly isolated from whole blood in CPT; BCT-CPT, PBMC isolated from BCT-preserved whole blood and transferred into CPT. Error bars represent ± two standard error of the mean for each side. PBMC, peripheral blood mononuclear cell. CPT, Cell Preparation Tubes. BCT, Cell-Free DNA Blood Collection Tubes.

### Clinicopathological Characteristics of ESCC Patients

Five ESCC cases were divided into low and high tumor burden groups. By endoscopy, EC_1 and EC_5 showed superficial primary tumors with invasion to the submucosal layer (Figure 5 A, B). Because metastatic lesions were not detected by CT scan, both cases were clinically diagnosed as Stage I. Thoracoscopic subtotal esophagectomy with gastric tube reconstruction and lymphadenectomy were performed for both cases. Pathological examinations of resected specimens revealed that EC_1 had submucosal invasion and three regional lymph node metastases. Therefore, EC_1 was diagnosed as pathological stage IIIA, even though this case had a low tumor burden. EC_5 was diagnosed as stage I with submucosal invasion with no metastases. Both cases showed no recurrence during a three year follow-up.

**Figure 5.**
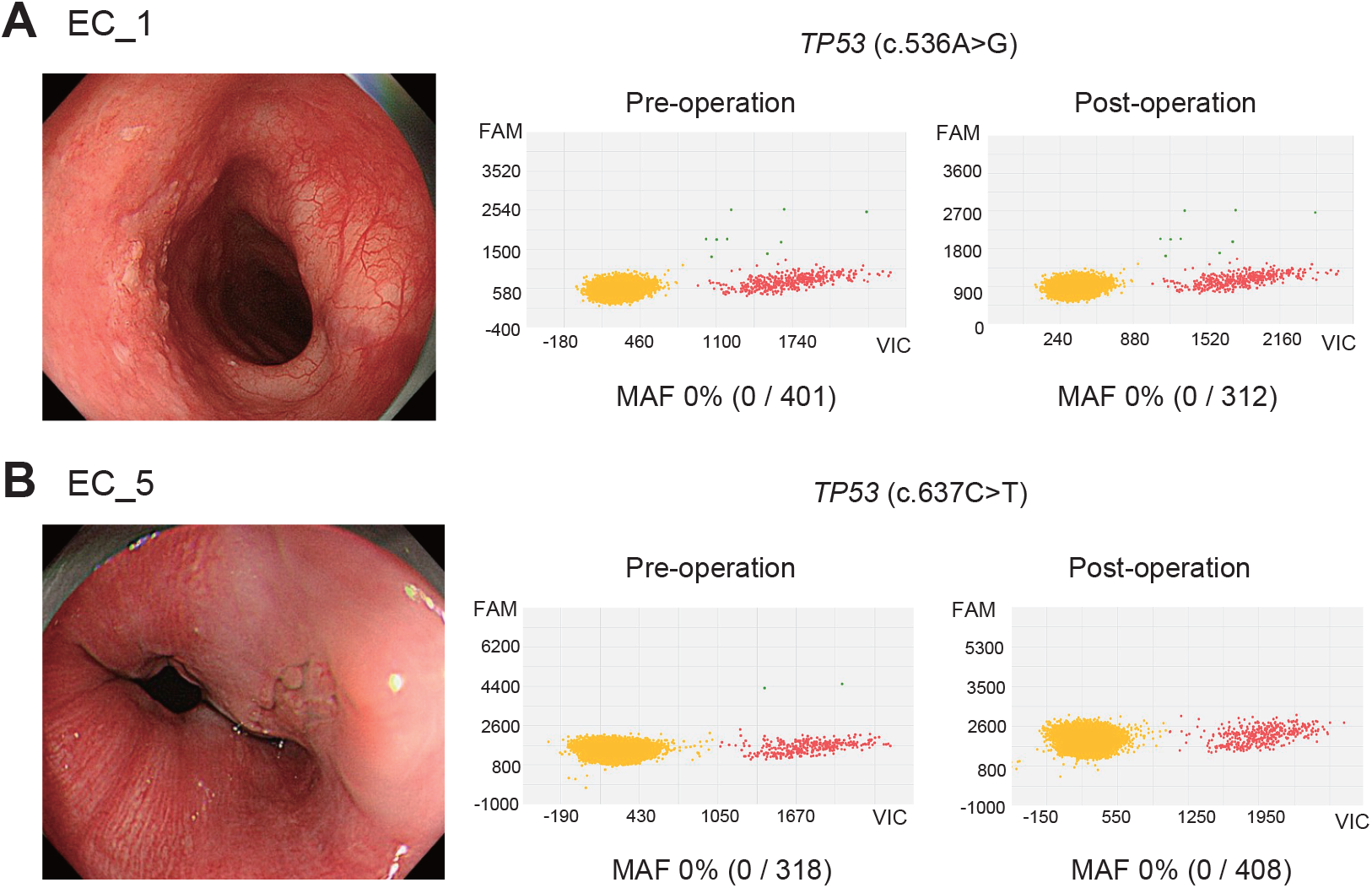
Quantitative tumor burden monitoring by ctDNA detection in low tumor burden ESCC patients. Left panel, endoscopic image; right panel, scatter diagrams of dPCR analyses from pre-operation and post-operation plasma samples of EC_1 and EC_5 (**A** and **B**, respectively). Red dots indicate positive reactions for wild type probe whereas yellow dots indicate negative reaction for both wild type and mutant probes. dPCR, digital PCR. ctDNA, circulating tumor DNA. ESCC, esophageal squamous cell carcinoma. VAF, variant allele frequency. FAM, carboxyfluoscein.

EC_2, EC_3, and EC_4 had apparently higher tumor burden with primary and metastatic regions relative to EC_1 and EC_5. EC_2 had a primary tumor with adventitia invasion in the lower third of the esophagus and significant abdominal lymph node metastasis that had invaded the stomach. After chemotherapy, a marked tumor shrinkage was observed for this case (Figure 6A, bottom). Six cycles of chemotherapy followed by surgical resection were completed, and no recurrence was observed for three years. EC_3 had a primary tumor with adventitia invasion in the middle third of the esophagus, multiple abdominal lymph node metastases, and hematogenous metastasis in the left adrenal grand. Although the tumor size was temporarily reduced after two cycles of chemotherapy, the patient died of cancer progression 138 days after treatment initiation (Figure 6B). EC_4 had a cervical esophageal tumor that had invaded adjacent organ and exhibited a regional lymph node metastasis (Figure 6C). The 1st line chemotherapy treatment had no effect, so subsequent chemoradiotherapy and second-line chemotherapy were administered. The patient died of cancer progression 659 days after treatment initiation.

**Figure 6.**
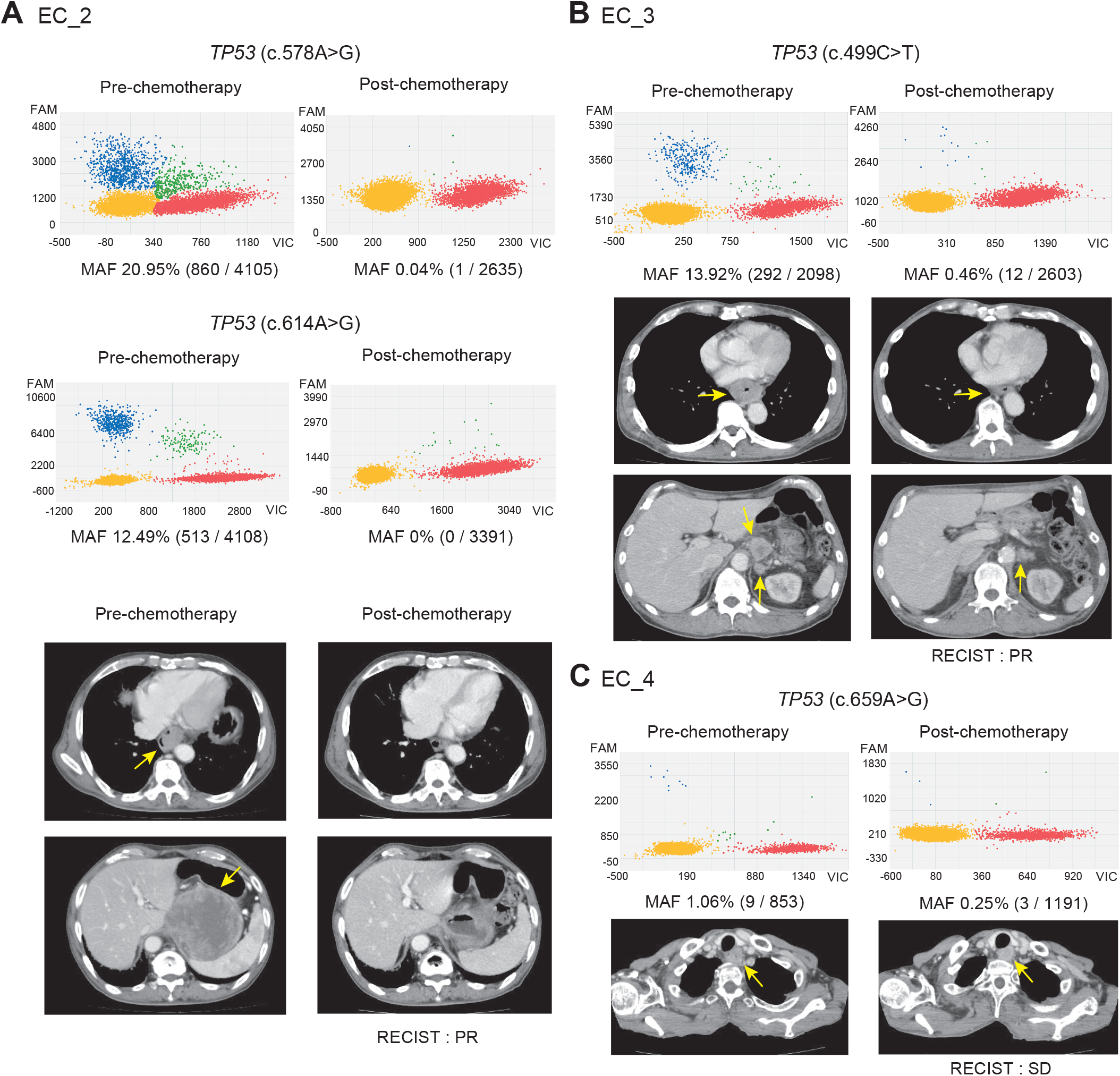
Quantitative tumor burden monitoring by ctDNA for advanced stage ESCC patients with high tumor burden. **A**, Scatter diagrams of dPCR analyses from pre- and post-chemotherapy plasma samples with two unique *TP53* mutations (upper four panels); and CT scan findings (lower four panels). **B**, Scatter diagrams of dPCR analyses from pre- and post-chemotherapy plasma samples with one unique *TP53* mutation (upper two panels); and CT scan findings (lower four panels). **C**, Scatter diagrams of dPCR analyses from pre- and post-chemotherapy plasma samples with one unique *TP53* mutation (upper two panels); and CT scan images (lower two panels). Blue and red dots indicate positive reactions for mutant and wild type dPCR probes, respectively. The indicated primary and metastatic tumors were visible by CT scan. ctDNA, circulating tumor DNA. ESCC, esophageal squamous cell carcinoma. dPCR, digital PCR. VAF, variant allele frequency. FAM, carboxyfluoscein. RECIST, response evaluation criteria in solid tumors. PR, partial response SD, stable disease.

### *TP53* Mutation Detection in Primary Tumor and Plasma DNA from ESCC Patients

The ctDNA levels and clinical tumor burden were evaluated for the five ESCC patients by *TP53* mutation detection. Sequencing analysis of all coding regions of *TP53* in five primary ESCC tissues revealed six non-synonymous mutations (Table 1, sequence data are available at DNA Data Bank Japan^26^, accession number JGAS00000000219). Each sample underwent an average of 50,397 sequencing reads after quality filtering. Average base coverage depth of targets in tumors, PBMCs, and total samples were 2273 (range: 1714-2613), 2099 (1941-2210), and 2186 (1714-2613), respectively. One missense and one nonsense mutation in exon 5 and three missense and one nonsense mutations in exon 6 were observed. The average VAF of the six mutations from the primary tumors was 36.4% (ranging from 13.4 to 70.3%). These mutations were targeted for ctDNA detection with plasma DNA. Blood samples were obtained in BCT followed by plasma separation and PBMC isolation in CPT. Plasma and PBMCs were separated within five days of WB preservation at room temperature. No significant somatic mutations from PBMC were detected in all five cases.

**Table 1.**
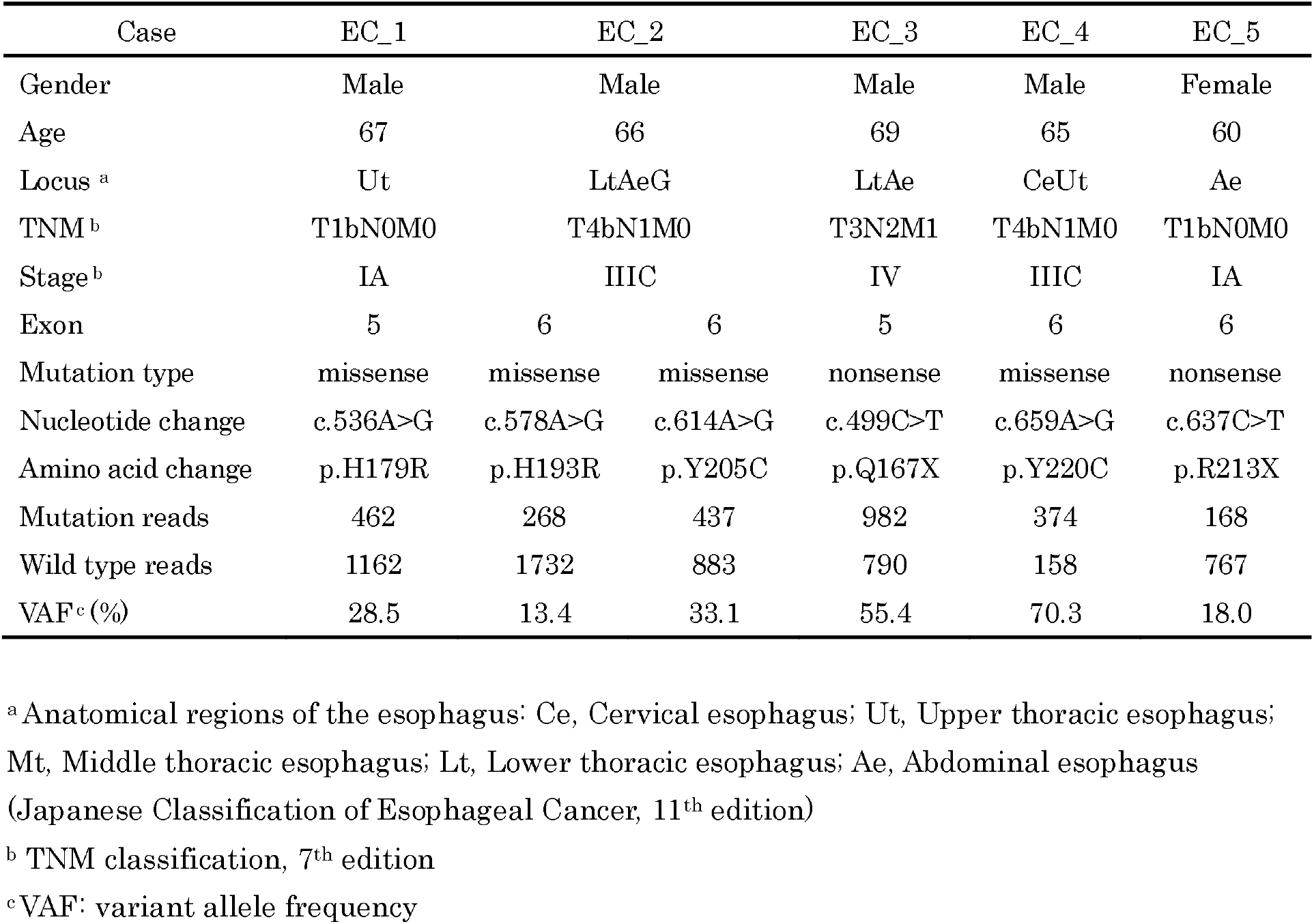
*TP53* mutation in primary ESCC tumors

Case-specific mutations in the corresponding plasma samples of pre- and post-treatment (i.e., surgery, triplet chemotherapy with cisplatin, 5-FU, and docetaxel) for ESCC patients were monitored using dPCR primer/probe sets designed for this study (primer/probe information may be provided upon request). In EC_1 and EC_5, post-surgical blood samples were collected at 14 days after esophagectomy. In EC_2, EC_3, and EC_4, post-chemotherapeutic blood samples were collected at three weeks after the first cycle of five-days of chemotherapy. Plasma samples from the two low tumor burden cases (EC_1 and EC_5) showed no detectable ctDNA, either pre- or post-surgery (Figure 5 A, B). Despite the substantial level of VAF in the primary tumors (28.4% and 17.7%, respectively), pre-operative undetectable ctDNA suggested that the tumor burden was low in these two cases. In contrast, the post-chemotherapy VAFs of ctDNA for the high tumor burden cases (EC_2, EC_3, and EC_4) were all decreased, consistent with the measurable tumor burden observed by CT scan. Notably, dPCR enabled detection of extremely low VAFs that were < 1% (Figure 6). Most of the VAFs for post-operative patients with curative intent were < 1%, which may not be a practical sequence depth for other exploratory techniques using a next-generation sequencing method such as exome sequencing. Although dPCR covers only mutations for which specific primer/probe sets are prepared for mutant and wild-type allele detection, our results indicate that dPCR could be a suitable technique for follow-up of post-treatment ESCC patients with high tumor burden. The case-specific dynamics of ctDNA in response to treatment are summarized in Table 2. All plasma DNA samples were obtained from BCT tubes and paired with germline DNA obtained from BCT-CPT-PBMC.

**Table 2.** ctDNA Dynamics Using Case-Specific TP53 Mutation in ESCC Patients

| Case | EC_1 | EC_2 |  | EC_3 | EC_4 | EC_5 |
| --- | --- | --- | --- | --- | --- | --- |
| *TP53* mutation | p.H179R | p.H193R | p.Y205C | p.Q167X | p.Y220C | p.R213X |
| Treatment | Surgery | DCF<sup>b</sup> |  | DCF | DCF | Surgery |
| Therapeutic effect of chemotherapy <sup>c</sup> | - | PR |  | PR | SD | - |
| Pre-treatment (rxns) |  |  |  |  |  |  |
| Mutation reactions | 0 | 860 | 513 | 292 | 9 | 0 |
| Wild type reactions | 401 | 4105 | 4108 | 2098 | 853 | 318 |
| VAF <sup>a</sup> (%) | 0 | 20.95 | 12.49 | 13.92 | 1.06 | 0 |
| Post-treatment (rxns) |  |  |  |  |  |  |
| Mutation reactions | 0 | 0 | 0 | 12 | 3 | 0 |
| Wild type reactions | 312 | 3343 | 3391 | 2603 | 1191 | 408 |
| VAF (%) | 0 | 0 | 0 | 0.46 | 0.25 | 0 |

<sup>a</sup> VAF, variant allele frequency<sup>b</sup> triplet chemotherapy with docetaxel, cisplatin, and 5-fluorouracil<sup>c</sup> PR, partial response; SD, stable disease (Response Evaluation Criteria in Solid Tumor, RECIST, v1.1)

## Discussion

Recent studies that compared characteristics of conventional blood collection tubes (e.g., BD Vacutainer K_2_EDTA tubes, K_3_EDTA tubes) demonstrated that BCT offers notable advantages for preservation of blood samples that will be used for cell-free DNA detection.^3, 4 27^ BCT prevents release of genomic DNA from nucleated cells into the blood and allows the samples to be stored for between four and 14 days before plasma DNA extraction.^3-5^ Our comparisons of BCT to CPT in this study demonstrated that the plasma layer in both tubes showed hemolysis. This observation suggests that the proprietary fixative in BCT specifically prevented nucleated cells from cytolysis, which is consistent with the known differences in fixation and permeabilization procedures for RBC and nucleated cells.^24^ To define the optimized practical conditions for our procedure, plasma DNA levels from six time points within 14 days of sample collection from healthy volunteers were validated in BCT and CPT. We observed no significant increase in plasma DNA or *LINE-1* copy number in BCT until 14 and 12 days, respectively, indicating that blood samples can be preserved for ctDNA detection for up to 12 days.

It should be noted that age-related clonal hematopoiesis are frequently found in the peripheral blood of healthy elderly individuals. It has been reported that 25% of non-hematologic cancer patients had at least one somatic mutations in their hematopoietic cells, whereas the mutations were not present in the corresponding tumor tissue.^28^ In practice, it has been confirmed that 8% of clinically reported “mutated genes” from a commercial ctDNA assay were in fact true clonal hematopoiesis events.^29^ Therefore, high-quality DNA extraction from PBMC or germline is necessary for cancer clinical sequencing, particularly for ctDNA analysis.

Wollison et al. demonstrated that fixative in BCT did not impact PBMC DNA quality by whole exome sequence and copy number microarray analyses.^30^ In their sample collection procedure, plasma was stored after centrifugation of collected blood followed by PBMC DNA extraction from remaining blood mixed with RBC and white blood cells using the Gentra Puregene Blood Kit (Qiagen, Hilden, Germany). According to the kit protocol, several hours are required after centrifugation and removal of plasma to complete PBMC DNA extraction. Our combinatory method using both BCT and CPT for a long-term preservation enables high quality DNA extraction of both plasma and PBMC within seven days at room temperature. We also demonstrated that PBMC isolation can be readily performed within seven days of collection by transferring whole blood WB samples stored in BCT to CPT. These results indicate that storage of WB in BCT maintains cellular membrane stability at room temperature for at least seven days. In practice, these procedures will facilitate ctDNA monitoring at centralized laboratories by providing enhanced shipping and handling feasibility across geographically broad areas.^3,4^

The target VAF for mutation detection in plasma by dPCR is generally 0.01-23% in most cancer patients.^37^ As the amount of plasma DNA is quite small (often less than 10 ng/mL plasma), the copy number of the mutated allele is often below the detection limit (i.e., one ng genomic DNA is equivalent to approximately ~333 copies of an allele, assuming that one bp is 618 g/mol and one genome copy is ~3.0 × 10^9^ bp). Due to these physiological characteristics, obtaining the necessary sequence depth to precisely detect such genuine rare mutations in plasma for exploratory sequencing by NGS is difficult. ^8, 24, 38, 39^ For the three ESCC patients with high tumor burden in this study, a decrease in VAF was confirmed by dPCR after chemotherapy treatment. Therefore, obtaining the mutation profile of primary tumors with sufficiently sensitive technologies is necessary to increase the usefulness of ctDNA detection in post-treatment cancer monitoring.^2, 8^ Particularly, post-treatment cancer patient follow-up requires multiple time measurements of mutations that are unique to the patient. Our proposed procedure would thus be one of the most practical blood collection approaches for daily practice.

In summary, our results indicate the following: (i) blood samples can be preserved for a week long-term in daily practice; (ii) our combinatory tube method can preserve plasma and PBMC with simple centrifugation; and (iii) the method yields DNA that can be reliably used for tumor-burden monitoring in advanced ESCC patients. The technical procedures that we established in this study will potentially be an important component of ctDNA-associated diagnosis in daily clinical practice.

## Data Availability

Sequence data were deposited in the DNA Data Bank Japan (Accession number JGAS00000000219).

## Acknowledgements

We thank all study participants, as well as the nursing and laboratory staff of Iwate Medical University Hospital. We also thank the healthy volunteers who provided blood samples. This study was supported by Keiryokai Collaborative Research Grant [#131, #136] and Grant-in-Aid for Scientific Research KAKENHI [JP16H01578, JP16K19951, JP16K19952, JP17K10605, and JP16H06279].

